# Feasibility Study of Mitigation and Suppression Strategies for Controlling COVID-19 Outbreaks in London and Wuhan

**DOI:** 10.1101/2020.04.01.20043794

**Authors:** Po Yang, Jun Qi, Shuhao Zhang, Xulong wang, Gaoshan Bi, Yun Yang, Bin Sheng, Geng Yang

**Author notes:** { } (Po Yang).

## Abstract

Recent outbreaks of coronavirus disease 2019 (COVID-19) has led a global pandemic cross the world. Most countries took two main interventions: suppression like immediate lockdown cities at epicentre or mitigation that slows down but not stopping epidemic for reducing peak healthcare demand. Both strategies have their apparent merits and limitations; it becomes extremely hard to conduct one intervention as the most feasible way to all countries. Targeting at this problem, this paper conducted a feasibility study by defining a mathematical model named SEMCR that can access effectiveness of mitigation, suppression and hybrid interventions for controlling COVID-19 outbreaks in London and Wuhan. It extended traditional SEIR (Susceptible-Exposed-Infectious-Recovered) model by adding two key features: a direct connection between Exposed and Recovered populations, and separating infections into mild and critical cases. It defined parameters to classify two stages of COVID-19 control: active contain by isolation of cases and contacts, passive contain by suppression or mitigation. The model was fitted and evaluated with public dataset containing daily number of confirmed active cases including Wuhan and London during January, 2020 and March 2020. The simulated results showed that 1) Immediate suppression taken in Wuhan significantly reduced the total exposed and infectious populations to 119610, but it has to be consistently maintained at least 90 days (by the middle of April 2020). Its success heavily relied on sufficiently external support from other places of China. This mode were not suitable to other countries that have no sufficient health resources. 2) In London, it is possible to take a hybrid intervention of suppression and mitigation for every 2 or 3 weeks over a longer period to balance the total infections and economic loss. While the total infectious populations in this scenario would be possibly 2 times than the one taking suppression, economic loss and recovery of London would be less affected. 3) Both in Wuhan and London cases, one important issue of fitting practical data was that there were a large portion (probably 62.9% in Wuhan) of self-recovered populations that were asymptomatic or mild symptomatic. These people might think they have been healthy at home and did not go to hospital for COVID-19 tests. Early release of intervention intensity potentially increased a risk of the second outbreak. One limitation of our model was that our prediction of infections and deaths depended on a parameter estimation of intervention intensity that presented by average-number contacts with susceptible individuals as infectious individuals in a certain region. It assumed that each intervention had equivalent effects on the reproduction number R in different regions over time. Practical effectiveness of implementing intervention intensity might be varied with respect to cultures or other issues of certain county.

## 1 INTRODUCTION

Throughout human history, Infectious diseases (ID), also known as transmissible diseases or communicable diseases, are considered as serious threats to global public health and economics [1]. From the 1918 influenza pandemic in Spain resulting in nearly 50 million deaths in 1920s, to recent ongoing global outbreaks of coronavirus disease 2019 (COVID-19) killing over 11 thousands people in all over the world [2], infectious disease is a leading contributor to significant mortality and causes huge losses to society as well as personal family burden. Among a variety of factors leading to emergence and outbreaks of ID, the key issues are population density and human mobility where in these cities with developed transportation systems, pathogens can be spread to large geographic space within a short period of time. For instance, the ongoing global epidemic outbreak of COVID-19 has spread to at least 146 countries and territories on 6 continents in 2 months. In order to give an accurate prediction of outbreaks, many researchers have been working in traditional ID propagation models [3-7] like SIR, SEIR,.et.al, for understanding COVID-19 transmission with human mobility and predicting outbreak process of epidemics. Meanwhile, as realizing a long period of this battle against COVID-19, many of them recently focus on intervention strategies [8-10] that can balance a trade-off between limited human mobility and potential economic loss in COVID-19 control. It poses an important research area that explores how and when to take what level of interventions in light of multiple natures and capabilities of countries.

In traditional compartmental models paradigm in epidemiology, SIR (Susceptible-Infectious-Recovered) [3] and SEIR (Susceptible –Exposure-Infectious-Recovered) [4] are two popular approaches to simulate and predict how infectious disease is transmitted from human to human. These two models have defined several variables that represent the number of people in each compartment at a particular time. As implied by the variable function of time, these models are dynamic to reflect the changes and fluctuations of these numbers in each compartment over time. For COVID-19 control in Wuhan, Zhong, et.al [6] introduced a modified SEIR model in prediction of the epidemics trend of COVID-19 in China, where the results showed that under strong suppression of “lockdown Hubei”, the epidemic of COVID-19 in China would achieve peak by late February and gradually decline by the end of April 2020. Some other extended models [8] [12] are also proposed for predicting the epidemics of COVID-19 in Wuhan and give some similar forecasts. While above methods demonstrate good performance in prediction of COVID-19 outbreak by taking strong public intervention, also named as suppression strategy [13] that aims to reverse epidemic growth, one important challenge is that taking suppression strategy only is to treat disease controls as single-objective optimisation of reducing the overall infectious populations as soon as possible, and require strategic consistency in a long term. In real-world, taking public health intervention strategies is actually a multiple-objective optimisation problem including economic loss and society impacts. Thus, most countries have taken different intervention strategies, like enhanced surveillance and isolation to affected individuals in Singapore [14], four-stage response plan of the UK [15], mitigation approaches [13] and even multiple interventions taken in many EU countries. Due to the fact that standalone intervention strategy has apparent merits and limitations, it becomes highly necessary to study the feasibility of intervention strategies to certain country in light of its multiple natures and capabilities.

Targeting at this problem, this paper conducts a feasibility study that analyses and compares mitigation and suppression intervention strategies for controlling COVID-19 outbreaks in Wuhan and London. As shown in Fig.1 with Wuhan as a simulated case using data from [6], we demonstrated performance of taking different intervention strategies: a) No interventions: the peak of daily infections would be up to 2.1 million, but will be completed in 150 days. The epidemics lasts a relatively shorter period of 140 - 150 days, but lead to more death. b) Contain phase: taking 90% effectiveness of surveillance and isolation from the 2^nd^ day of confirmed case potentially enables controlling a new outbreak of COVID-19, but it needs to be maintained over 300 days. c). Suppression intervention from the 32^nd^ day: the peak of daily infections greatly reduced to 16 thousand, but it had to be followed at least 200 days. Nearly 3 months suppression may potentially lead to economic loss even crisis. d). Mitigation intervention from the 32^nd^ day: the peak of daily infectious populations increased to 27.7 thousand, but the period of maintenance extended to 150 days. It implied there would be growing death but less economic loss compared to suppression. e). Hybrid intervention of taking both suppression and intervention every 2 weeks: the epidemics of COVID-19 appeared a long-term multimodal trend where the peaks of daily infectious populations were within a range of 40-60 thousand. This might lead to less daily critical cases and offer more time to hospital for releasing their resources.

**Figure 1:**
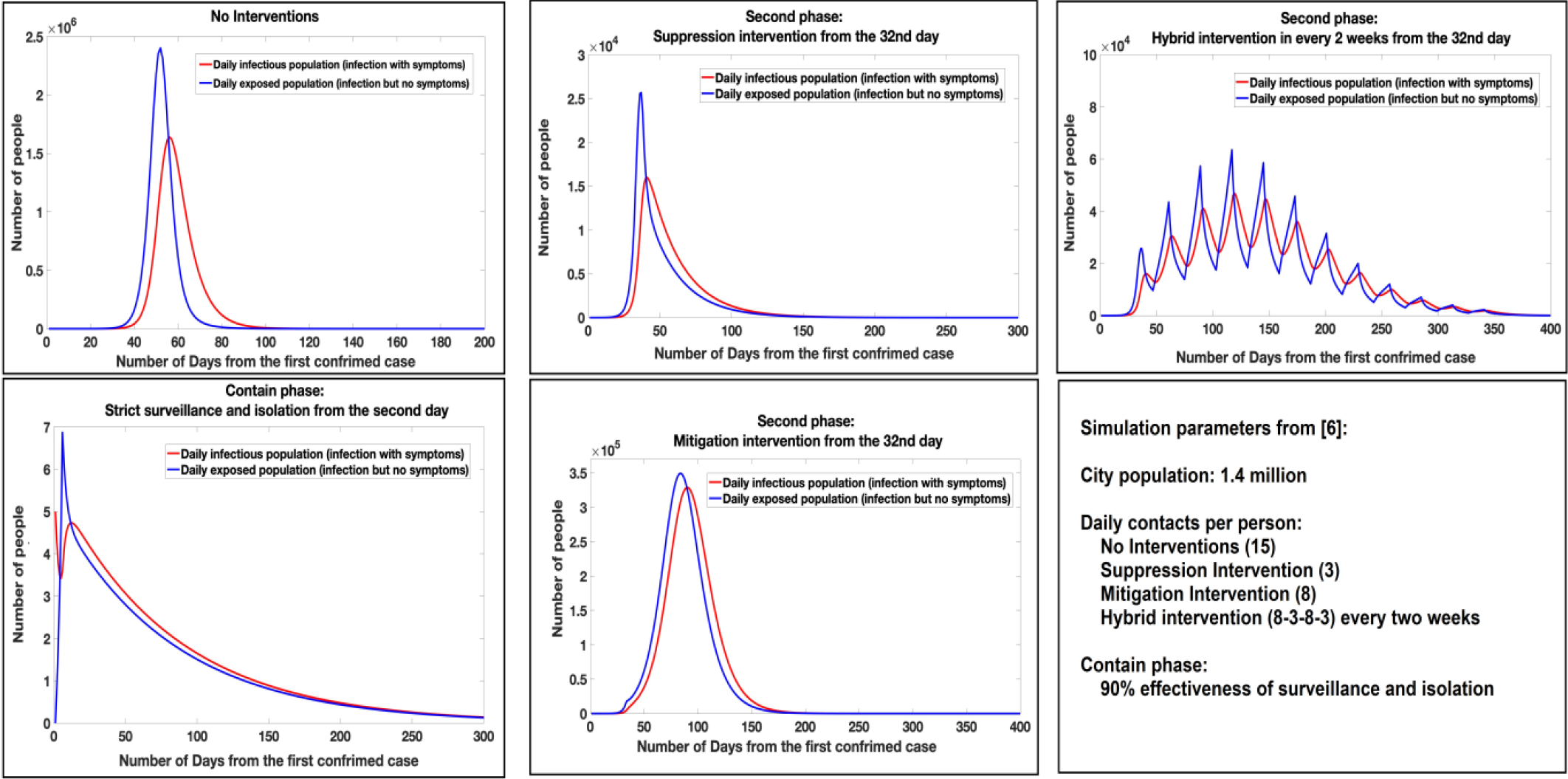
**Illustration of controlling Wuhan COVID-19 outbreaks by taking different intervention strategies with parameters (City populations: 1.4 million; daily contacts per person: No Interventions (15), Suppression Intervention (3), Mitigation Intervention (8), and Hybrid intervention (8-3-8-3) every two weeks; Contain phase (98% effectiveness of surveillance and isolation))**.

Above analysis demonstrates the complexity of controlling COVID-19 outbreaks that how and when to take what level of interventions. In this paper, we proposed a mathematical model: SEMCR to study this problem. The model extended traditional SEIR (Susceptible-Exposed-Infectious-Recovered) model [3] [4] by adding one important fact: there has been a direct link between Exposed and Recovered population. Then, it defined parameters to classify two stages of COVID control: active contain by isolation of cases and contacts, passive contain by suppression or mitigation. The model was fitted and evaluated with public dataset containing daily number of confirmed active cases including Wuhan, London, Hubei province and the UK during January, 2020 and March 2020. For each point, we design and set up experimental protocols for comparison and exploration, highlighting following multi-fold contributions:

- Immediate suppression taken in Wuhan significantly reduced the total exposed and infectious populations, but it has to be consistently maintained at least 90 days (by the middle of April 2020). Its success heavily relied on sufficiently external support from other places of China. This mode were not suitable to other countries that have no sufficient resources.
- In London, it is possible to take a hybrid intervention of suppression and mitigation for every 2 or 3 weeks over a longer period to balance the total infections and economic loss. While the total infectious populations in this scenario would be possibly 2 times than the one taking suppression, economic loss and recovery of London would be less affected.
- Both in Wuhan and London, one important issue of fitting practical data was that there were a large portion (at least 42.5% in Wuhan) of self-recovered populations that were asymptomatic or mild symptomatic. These people might think they have been healthy at home and did not go to hospital for COVID-19 tests. Early release of intervention intensity potentially increased a risk of the second outbreak.
- One limitation of our model was that our prediction of infections and deaths depended on a parameter estimation of intervention intensity that presented by average-number contacts with susceptible individuals as infectious individuals in a certain region. It assumed that each intervention had equivalent effects on the reproduction number R in different regions over time. Practical effectiveness of implementing intervention intensity might be varied with respect to culture or other issues of certain county.

The remainder of this paper is arranged as follows. Section 2 introduces the model for controlling COVID-19 outbreak. In the Section 3, the materials and implementation of experiment are reported. Section 4 provides detailed experimental evaluation and discussion. The conclusion and future directions are given in Section 5.

## 2 METHODOLOGY

### 2.1 Problem formulation of COVID-19 outbreak

We modified a SEIR model to account for a dynamic Susceptible [S], Exposed [E], Infectious [I] and Recovered [R] or Dead [D] population’s state by extending two components: Mild [M] cases and Critical [C] cases. The modified modal is shown in Fig.2. Here, we assume that susceptible population *E* represents susceptible population of a certain region; and *β* represents effectiveness of intervention (strict isolation) in contain phase. If effectiveness of intervention in contain phase is not sufficiently strong, susceptible individuals may contract the disease with a given rate when in contact with a portion of exposed population (asymptomatic but infectious) *E*. After an incubation period, the exposed individuals become the infectious population *I* (symptomatic) at a ratio α_1_. Notably, infectious population starts from mild cases *M* to critical cases *C* at a ratio *α*_*2*_. Finally, a portion *d* of critical cases lead to deaths; the rest of infectious population will be recovered.

**Figure 2:**
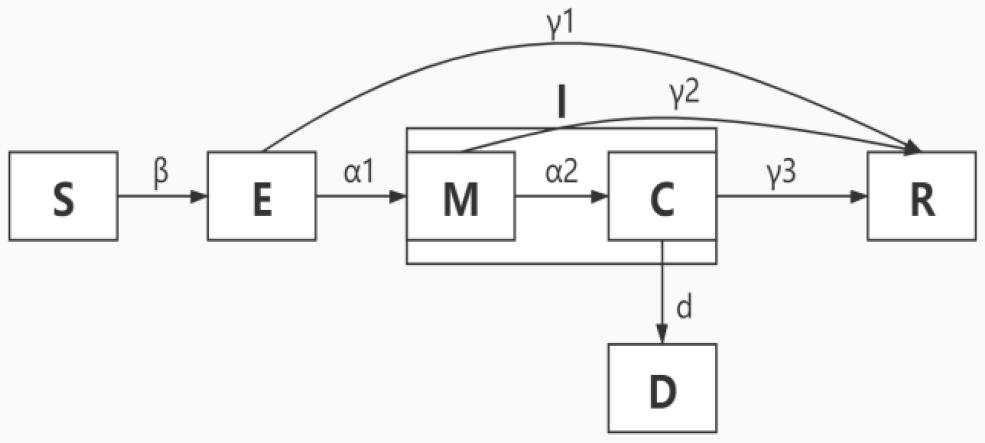
**Extended SEMCR model structure: The population is divided into the following six classes: susceptible, exposed (and not yet symptomatic), infectious (symptomatic), mild (mild or moderate symptom), critical (sever symptom), death and recovered (ie, isolated, recovered, or otherwise non-infectious)**.

There are two enhanced features in our model in comparison to popular SEIR models [6] [8] [12]. The first one is a straightforward relationship between Exposed and Recovered population. We find that in the early outbreaks of COVID-19, some portion of exposed people may have no obvious symptoms or only develop as mild cases, but they cannot get a test due to lack of testing kits. This group of populations might be self-recovered in some days, but will not realize they were infected. The second feature in our model is that we separate infectious population into mild and critical cases in light of their symptoms. It mainly concerns a phenomena of relatively higher mortality rate in the early outbreak of COVID-19 in Wuhan after taking immediate suppression intervention on 23^rd^ Jan 2020. Most likely, suppression intervention has led to fear in Wuhan citizens and chaos in hospital systems, as a result of delaying rescue of many critical cases.

Introducing above two features, it is helpful for evaluating real effects of different interventions. If we assumed the overall population of a certain region is *N*, the number of days is *t*, the dynamic transmissions of each components of our model are defined as follow:

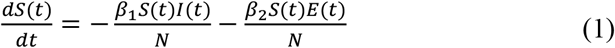

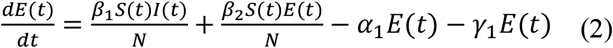

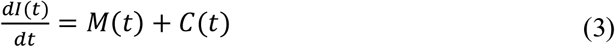

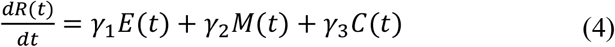

Regarding Mild cases, Critical cases and Death, the dynamic transmission is as below:

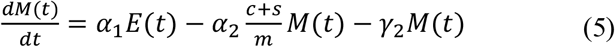

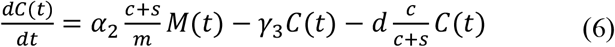

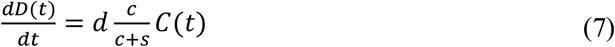

Lastly, we define a benchmark in SEMCR model to reflect the strength of intervention over time, as M_t_. It is presented by average number of contacts per person per day in a region.

### 2.2 Implementation of dynamic transmission of SEMCR

In practical cases, it needs to estimate the defined parameters including *α*_1_, *α*_2_,β, and *γ*_1_, *γ*_2_, *γ*_3_, b, where β is the product of the people exposed to each day by confirmed infected people (k) and the probability of transmission (b) when exposed (i.e., β= kb) and σ is the incubation rate which is the rate of latent individuals becoming symptomatic (average duration of incubation is 1/*α*_1_). According to recent report [8], the incubation period of COVID-19 was reported to be between 2 to 14 days, we chose the midpoint of 7 days. *γ* is the average rate of recovery or death in infected populations. Using epidemic data from [6], we used SEMCR model to determine the probability of transmission (b) which was used to derive β and the probability of recovery or death (*γ*). The number of people who stay susceptible in each region was similar to that of its total resident population. Other transmission parameters were estimated with early prediction of Hubei cases in [6] on January 23 2020 using Monte Carlo simulation, as shown in the Table.1

**Table 1:** Parameters estimation in SEMCR model.

| Name | Representation | Value |
| --- | --- | --- |
| $N$ | Total number of population in a region | N/A |
| $\beta_1$ | Transmission rate for the I to S | 0.157 |
| $\beta_2$ | Transmission rate for the E to S | 0.787 |
| $\alpha_1$ | Incubation period | 6 |
| $\alpha_2$ | Incubation period from M to C | 7 |
| $\gamma_1$ | Average period from E to R | 0.283 |
| $\gamma_2$ | Average period from M to R | 7 |
| $\gamma_3$ | Average period from C to R | 14 |
| $d$ | Average period from C to D | 28 |
| $m$ | Mild proportion | 0.8 |
| $s$ | Severe proportion | 0.138 |
| $c$ | Critical proportion | 0.061 |
| $M_t$ | Intervention intensity | 3-15 |

Notably, as for the strength of intervention M, it was related to the population density in a region. We used a benchmark reported in [6] that assumes Hubei province with no intervention as M = 15, and after suppression intervention, M reduced to 3. When applying SEMCR model into other simulated cases, M was initialized according to the population density and human mobility in these places. Also, after taking any kind of interventions, the change of M would follow a reasonable decline or increase over few days, not immediately occur at the second day.

Following previous assumptions, the implementation of dynamic transmission of SEMCR model follows steps as below:

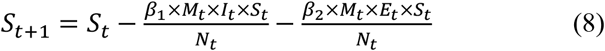

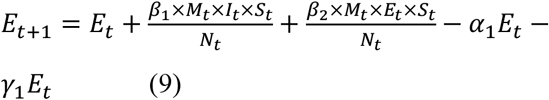

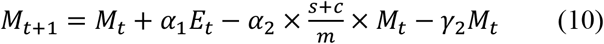

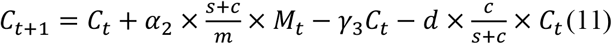

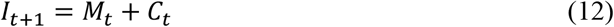

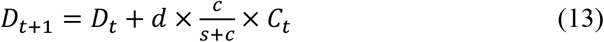

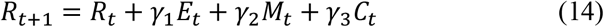

### 2.3 Model evaluation protocol

In order to utilise our proposed SEMCR model into practical cases, we design an evaluation protocol to access multiple effects of taking different intervention strategies to control outbreak of COVID-19 in 4 typical cases, including Hubei province, Wuhan city, the UK and London, as shown in Fig.3.

**Figure 3:**
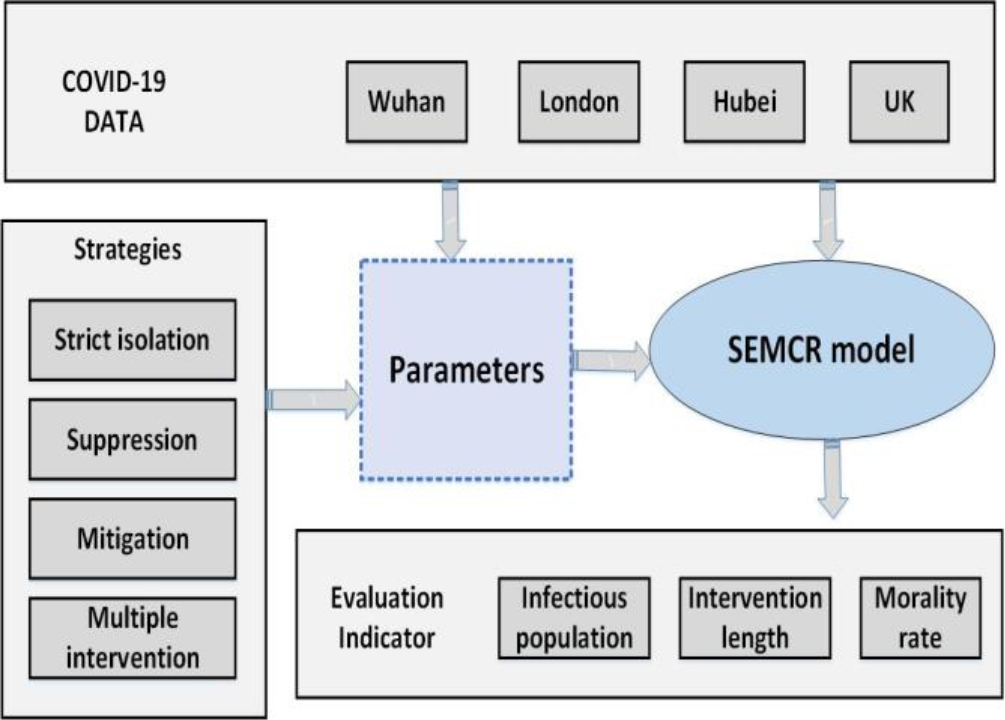
SEMCR model evaluation protocol.

The first stage is initial parameters estimation using COVID-19 data from four cases: Hubei, Wuhan, UK and London. In this stage, a preliminary qualitative assessment of each case is performed, by comparing their similarity and dissimilarity on area, transportation, population density, migration flows, date of the first confirmed case, etc. We would determine value of initial parameters in the SEMCR including N, M_t_, and date of the first confirmed case. Notably, in Wuhan, the date of the first confirmed case is not officially released. The work [8] estimates the first confirmed case is by the end of Nov 2019; and Zhong [11] points out the first confirmed case in Wuhan is on 23^nd^ December 2019. Here, we take the same settings of first confirmed case on 23^nd^ December 2019.

The next step is to estimate and normalize other parameters in the model. Assuming that COVID-19 has similar transmission ratio and incubation rate in all four cases, we use parameter values fitted from [11], where incubation rate is 1/7; the rate of transmission for the I to S is 0.157; the rate of transmission for the E to S is 0.787. As for estimation of other parameters, we follow the COVID-19 official report from WHO [15], including the proportion of Mild, Severe and Critical cases, the probability of death, etc.

Thirdly, how to take intervention strategies needs to be evaluated by tuning parameters in SEMCR model. The key tuning operation is to adjust the level of M_t_ over a period. For instance, we assume that no intervention strategies result in unaltered internal mobility of a region, taking suppression strategy in Wuhan means a reduction of M to 3. But in other cases with larger area, it is extremely difficult to take a complete suppression strategy. So the reduction of M will be relatively adjusted to 4 or 5.

Final stage, we perform quantitative analysis of effectiveness of different intervention strategies, including: strict surveillance and isolation, suppression strategy, mitigation strategy, and multiple intervention. The evaluation metric of cross-validation is employed to evaluate the performance of COVID-19 progression model. The final two evaluation indicators are the length of intervention and the peak time. The length of intervention is calculated due the date that confirmed cases are nearly clear to zero.

### 2.4 Data collection

The most recent epidemiological data based on daily COVID-19 outbreak numbers reported by the National Health Commission of China were retrieved. Also regarding the daily update from world meter, we record the number of confirmed cases and death each day in four cases. In order to simulate four cases, we require the exact confirmed cases in the first 2-5 days to initialise parameters of our model.

## 3 EXPERIMENTS

### 3.1 Evaluation of cases with no interventions

We simulated four cases that predict COVID-19 outbreaks without taking any interventions. The initial populations were given as London (9.3 million), Wuhan (14.18 million), UK (66.49 million) and Hubei (58.9 million). The parameter M representing average number of contacts per person per day was given as 15 to London, Wuhan and Hubei; 12 to the UK. The simulation results were given in Figure.4.

**Figure 4:**
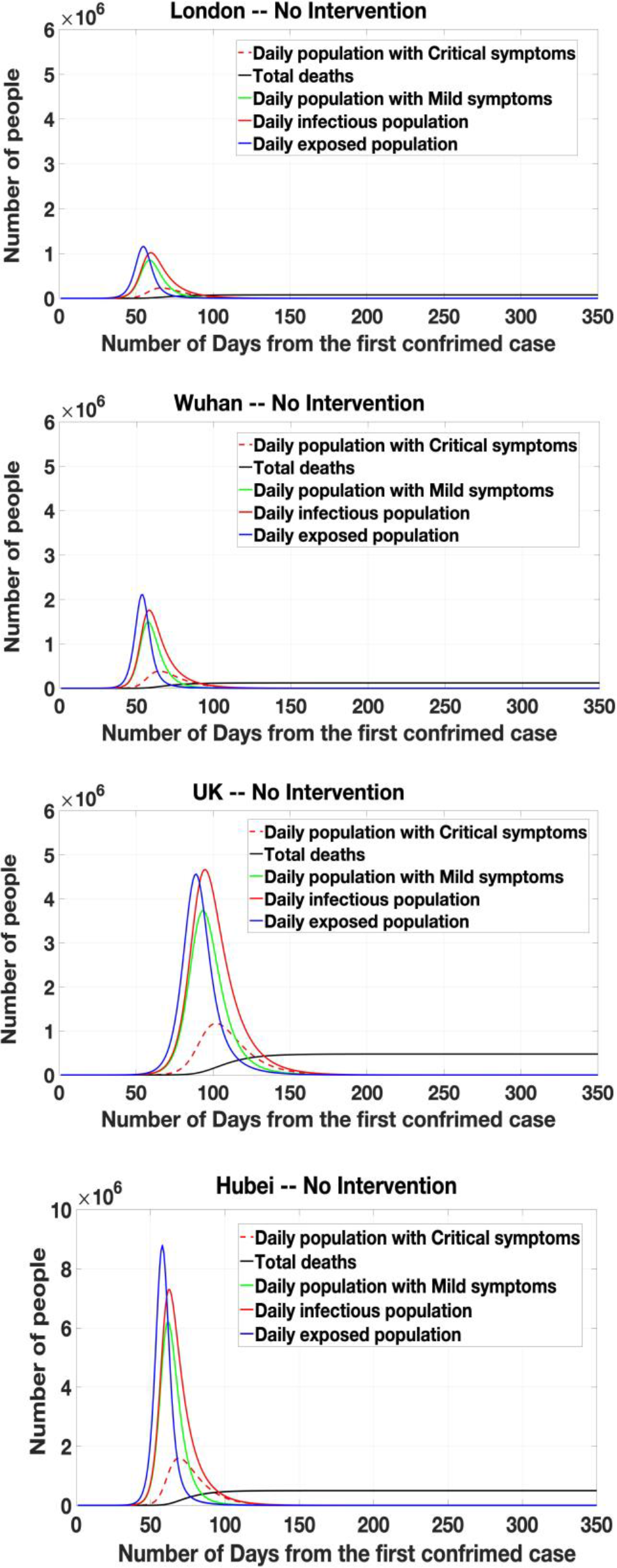
Four cases with no interventions.

The results showed that in the peak time, there would be up to 1.16 million, 210 thousand, 450 thousand and 900 thousand Exposed population (infection but no symptoms) at London, Wuhan, UK and Hubei. This implied that: 1) the total infectious population of these four cases would be 7.76 million in London, 12.27 million in Wuhan, 50.95 million in Hubei and 48.46 million in the UK. 2) The total death of these four cases would be 761 thousand in London, 120 thousand in Wuhan, 493 thousand in Hubei and 475 thousand in the UK. It equalled to about over 80% of total population of each region will be infectious, with the morality rate nearly 1%. It showed that without intervention, the outbreak of COVID-19 would lead to huge infections and deaths. The main reason was that COVID-19 was estimated as relatively high production number R_0_ up to 3 [14], where the transmission ratio β_2_ from Susceptible to Exposed is up to 78%. Thus, in some regions with high migration and dense population, it would easily lead to an outbreak.

But, one notable issue was that the initialisation of parameter M seems to impact on the occurrence of peak time and the length of overall period. In the cases of London, Wuhan and Hubei with M = 15, their peak date were all roughly on the 60^th^ day from the date of first confirmed case; but UK with M = 12, their peak time was delayed to the 100^th^ day from the date of first confirmed case. That meant to regions with similar total population, low population density potentially reduced overall infectious population, but delayed the peak time of outbreak as a result of longer period.

### 3.2 Effectiveness of surveillance and isolation

As for the strategy of taking surveillance and isolation intervention in the contain phase, recent study [15] developed a stochastic transmission model parameterised to the COVID-19 outbreak. It proved that highly effective contact tracing and case isolation can control a new outbreak of COVID-19 within 3 months, where for a production number R_0_ of 3.5, more than 90% contacts had to be traced. We transferred this finding as a tuning parameter β_2_ to evaluate if the outbreak of COVID-19 could be controlled.

Considering that 98% of contacts had to be traced, it implied that the surveillance and isolation was effective to scale down the group of contacts. In other words, we simulated a situation that from the second day of receiving confirmed case, only 5% of the overall population would be possible to contact the infectious ones. Then, the β_2_ = 0.05. The results were shown in the Fig.5. The results show that in all four cases, the outbreaks of COVID-19 were successfully controlled, and had no peak time. There would be only 44, 56, 42 and 120 people that were infection and finally recovered at London, Wuhan, UK and Hubei. The overall period of COVID outbreak was less than 100 days. This finding was as similiar as the proof in [15].

**Figure 5:**
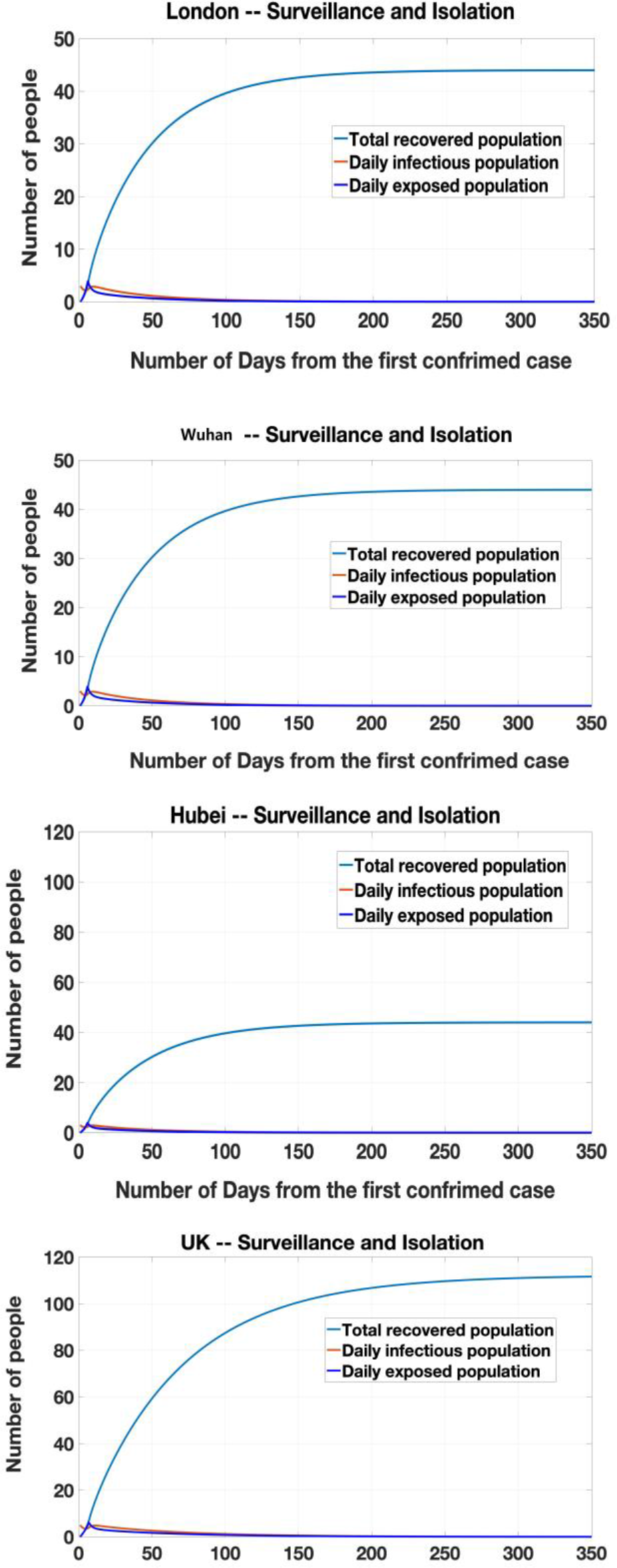
Four cases by taking surveillance and isolation in the contain phase.

Notably, the simulated situation of Wuhan, London and Hubei were same, it was mainly because we initialised the same value of confirmed cases as 3, and M = 15 in all cases. When taking highly effective surveillance and isolation, the transmission of COVID-19 was limited and controlled within a small group of population. The population difference would not affect the total infectious ones. But in the UK case with M = 12, low population density limited the effectiveness of surveillance and isolation, as a result of more infectious population. It implied another fact that to these countries with low population density, it was challenging to take high quality of intervention like surveillance and isolation.

Then, we evaluated when the outbreak of COVID-19 could occur by adjusting the value of parameter β_2_ in the UK case. As shown in Table.2, we recorded the total recovered population, and if there would be a peak of outbreak. As the increased β_2_, the total recovered population was dramatically increased and generated a peak as β_2_ = 0.075. That meant that if we cannot guarantee at least 92.5% of UK population being not contacted by infectious people, it would be indispensable to have an outbreak of COVID-19. If the isolation intervention was less effective; the more population would be infectious; and the peak day was brought forwarded. In the early stage, taking high effective surveillance and isolation in any regions is necessary to avoid the outbreak of COVID-19.

**Table 2:** Parameter tuning in the UK case.

| Value | Total recovered population | Peak | Peak Day |
| --- | --- | --- | --- |
| 0.055 | 190 | N | N |
| 0.060 | 1100 | N | N |
| 0.065 | 2742 | N | N |
| 0.070 | 130K | N | N |
| 0.075 | 1.90M | Y | 240 |
| 0.080 | 3.50M | Y | 230 |
| 0.090 | 4M | Y | 180 |

### 3.3 Effectiveness of Suppression Intervention

The suppression strategy was recognised as the most effective solution to reduce the infectious population, where it was taken in Wuhan on 23^rd^ January 2020. In the work [11], Zhong has reported that taking suppression strategy has successfully limits the overall infectious population in Hubei on 22^nd^ February 2020 to 50K. And if the suppression strategy was taken one week earlier, this figure would reduce to 18K. Thus, we would like to simulate the situation of taking suppression strategy in London. The parameter M was given as 15 to 4 in London, in comparison to Wuhan from 15 to 3. The simulation results were given in Figure.6.

The results showed that taking suppression strategy, the cases in Wuhan and London appeared a similar trend that daily exposed and infectious population would be greatly reduced. The outbreak of COVID-19 was controlled by the 100^th^ days, and can be nearly ended by the 150^th^ days. The difference was that the daily infectious population of London was nearly double to the ones in Wuhan. It was probably because we simulated the date of taking suppression strategy in Wuhan (the 32^nd^ day) is 3 days earlier than London (the 35^th^ day). Another possible reason was that considering the impact of culture difference, the value of M was only limited to 4 in London, but not as lower as 3 in Wuhan. In fact, the suppression in Wuhan was actually applied to limit mobility in community level with very high intensity, which was hard to be followed by London.

We considered another two situations of taking suppression intervention 1 week earlier or 1 week later in London, as shown in Fig.6. The results showed that the overall infectious population would be greatly reduced to 36 thousand if taking actions one week earlier; oppositely it would increase to 1.414 million infection if taking actions one week later.

**Figure 6:**
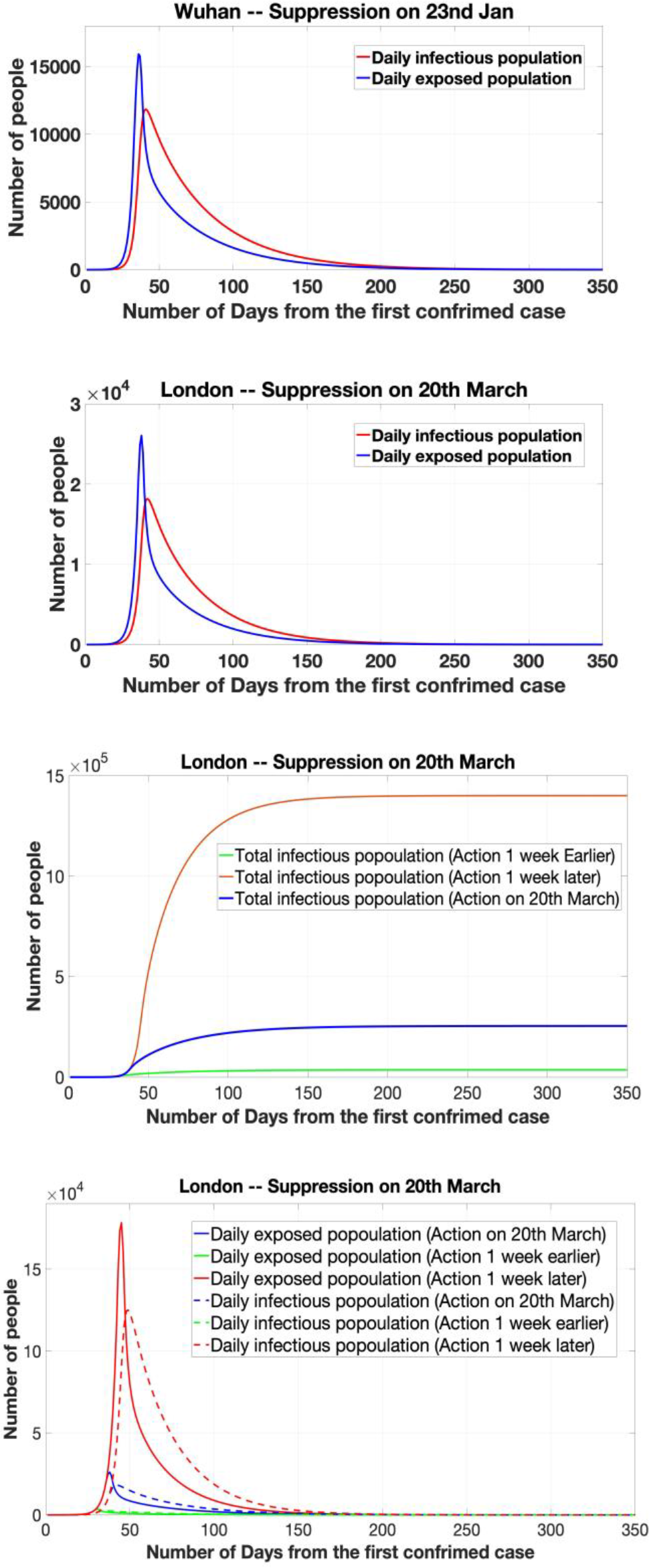
Wuhan and London by taking suppression intervention.

Another two important issues of taking suppression strategy were intervention length and the peak time. In order to effectively control the outbreak, the intervention length of taking actions at above three timing points all required at least 100 days. It potentially led to significant side effects to economic and society, including job loss, mental health, etc. Also, the peak time arrived earlier than its nature transmission if taking actions one week earlier. This might increase a shorter time to government for preparing sufficient resources, and causing more difficulties in control the outbreak of COVID-19.

### 3.4 Effectiveness of Mitigation Intervention

The mitigation strategy was recently highlighted by researchers from Imperial College [16] [17], where this strategy was initially taken by UK government. The aim of mitigation was to use other strategies to help individuals so that not to interrupt transmission completely, but to reduce the health impacts of an epidemic. In this cases, population immunity built up through the epidemic, leading to an eventual rapid decline in case numbers and transmission dropping to low levels. Thus, we simulated the situation of the UK by taking mitigation strategy with different level of strengths. The parameter M representing average number of contacts per person per day was given as 12 to the UK, and gradually reduced to 10 and 8 from the 32^th^ day of first confirmed case. The simulation results were given in Figure.7.

**Figure 7:**
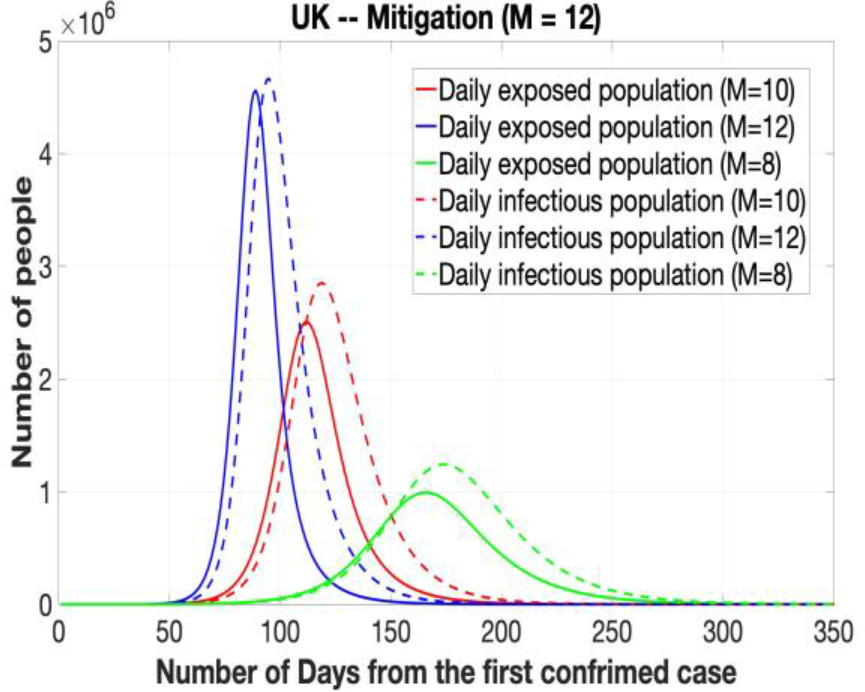
UK by taking mitigation intervention.

The results showed that taking mitigation strategy was effective to reduce daily infectious population, further lead to huge reduction of total infectious population. By reducing the parameter of M from 12 to 10 or 8, the peak of daily infectious population reduced from 4.5 million to 2.5 million or 1 million. On the other hand, the peak time of infectious population would be delayed as taking mitigation intervention, where the peak dates of infectious population at M = 12, 10 and 8 were roughly on the 80^th^, 110^th^ and 170^th^ days. The overall period of outbreak would be extended from 180 days to 200 or 250 days. Above simulated figures appeared a similar trend as findings from [17]. Taking mitigation intervention in the UK was capable of reducing the impact of an epidemic by flattening the curve, reducing peak incidence and overall death. While total infectious population may increase over a longer period, the final mortality ratio may be minimised at the end. But as similar as taking suppression strategy, the mitigation interventions need to remain in place for as much of the epidemic period as possible. However, the timing of introducing this mitigation intervention was important, where too early execution may allow transmission to return once they were lifted and sufficient “herd immunity” has not been developed.

### 3.5 Effectiveness of hybrid intervention

In terms of above discussion, the effectiveness of taking any one intervention (either suppression or mitigation) is likely to be limited. It is highly necessary to consider the possibility of taking multiple interventions to be combined to have a substantial impact on social and economic cost reduction. We simulated one simple situation of taking multiple strategy in London from the 35^th^ day, by giving a hybrid of suppression and mitigation strategies every 2 weeks. The M was given as a pattern of 6-4.5-3-4.5-6 in London, where it meant mitigation and suppression strategies were taken in an every 2 weeks roll. For minimizing side effects of taking interventions on human mobility, the application of first mitigation to reduce M from 12 to 6 spends 2 days. The simulation results were given in Figure.8.

**Figure 8:**
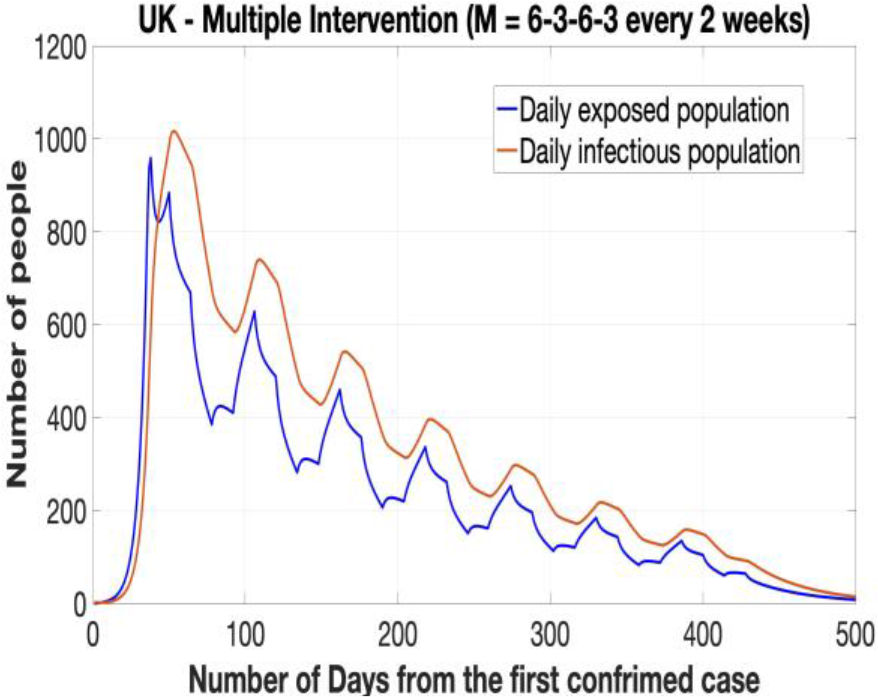
UK by taking hybrid intervention.

The results showed that the epidemic appeared a multi-modal decline trend over 500 days. The first peak of infectious population occurred on the 53^th^ days with 1017 infections after taking suppression intervention to reduce M from 12 to 3. After two weeks, mitigation strategies were taken so that the second peak of infectious population raised up to 750 infections. The total infectious population was 53075 over 500 days; the deaths was limited to 520.

Apparently, taking multiple intervention in the UK is capable of reducing the impact of an epidemic by fluctuating the curve, reducing overall infections and death. While the total period will be extended, final mortality ratio may be minimised at the end. But the longer period of limiting human mobility might increase economic risks and reduce employment ratio. There will be plenty of choices to taking multiple interventions through adjusting the strength and length of intervention. The consequence possibility show a similar multimodal curve but with different peak incidence.

## 4 DISCUSSION AND LIMITATION

As we mentioned in section 1, the question on how and when to take what level of interventions to control an epidemic is highly challenging, particularly in light of multiple natures and capabilities of countries. In many cases, it is even hard to evaluate effectiveness until the end of epidemic, and there is always controversy on taking any one intervention. However, our findings contribute to several useful suggestions on controlling COVID-19 outbreak.

The first point is that highly effective surveillance and isolation strategy is necessary to control an epidemic in early stage. Ideally, if this strategy can be executed in excellent level, there will be no huge outbreak later on. The case in Wuhan is a sample that have not taken any effective surveillance and isolation in the early stage of COVID-19, resulting of huge difficulties on late interventions. Also, considering the area, transportation, migration flows and population density of a region, most countries cannot achieve excellent level of isolation in contain phase, probably only in fine or good level. The outbreak of an epidemic is inavoidable. But considering its low social and economic cost, this strategy is still a cost-effective option.

Secondly, the cases in Wuhan and London approve high effectiveness of suppression strategy to reduce the overall infection. Probably in the UK or similar countries, suppression will minimally require a combination of social distancing of the entire population, home isolation of cases and household quarantine of their family members. But its practical effectiveness is not possible to achieve as same as Wuhan, as the success of suppression strategy in Wuhan is based on locking down human mobility to community level and sufficient resource support from other cities or provinces in China. If there are no sufficient external support, it will be risky to take intensive suppression to entire country due to huge impacts on its economics. Also, taking such intensive intervention to control an epidemic will need to be maintained until vaccine released (up to 12 months or more). If intensive interventions are relaxed at any time points, the transmission will quickly rebound. This is more like a multi-modal curve when taking multi-intervention strategies.

Thirdly, we also find out that while COVID-19 is estimated as a high production rate (R = 3), experimental evaluation results show that in either Wuhan or London cases fitted with real data in the last 6 weeks, high percentage of exposed or infectious population (at least 42.% of infectious population in Wuhan) are actually self-recovered. These people may have no or mild symptoms but been not checked as confirmed cases. This is one important issue that Zhong’s SERI model [11] has ignored. It will answer the model [8] predicts practical infectious population in Wuhan that ten times over figures in [11]. Similarly, it could explain the estimation of practical morality ratio can be varied in [11] and [8].

Lastly, one limitation of our model is that its prediction of infections and deaths depends on a parameter estimation of intervention intensity that presented by average-number contacts with susceptible individuals as infectious individuals in a certain region. We assume that each intervention has the same effect on the reproduction number in different regions over time. The practical effectiveness of implementing intervention intensity might be varied with respect to cultures or many other issues of certain county. As implementing hybrid intervention, the policy needs to be specific and well-estimated at each day according to the number of confirmed cases, deaths, morality ratio, health resources, etc.

## 5 CONCLUSION

This paper conducts a feasibility study by defining a mathematical model named SEMCR that analyses and compares mitigation and suppression intervention strategies for controlling COVID-19 outbreaks in London and Wuhan Cases. The model was fitted and evaluated with public dataset containing daily number of confirmed active cases including Wuhan, London, Hubei province and the UK. The experimental findings show that the optimal timing of interventions differs between suppression and mitigation strategies, as well as depending on the definition of optimal. In future, our model could be extended to investigate how to optimise the timing and strength of intervention to reduce COVID-19 morality and healthcare demand in mobile application.

## Data Availability

The date in this manuscript are from public dataset, which has been referenced in the manuscript. All code required to reproduce the analysis is available online at: https://github.com/TurtleZZH/Feasibility-Study-of-Mitigation-and-Suppression-Intervention-Strategies-for-Controlling-COVID-19.git

## DATA AND CODE

All data and code required to reproduce the analysis are available online at: https://github.com/TurtleZZH/Feasibility-Study-of-Mitigation-and-Suppression-Intervention-Strategies-for-Controlling-COVID-19.git

